# Classification of Recurrence Status After Surgical Treatment of Chronic Subdural Hemorrhage – A Machine Learning Approach

**DOI:** 10.64898/2026.03.25.26349323

**Authors:** Hussam Hamou, Julius Kernbach, Hani Ridwan, Kimberley Fay-Rodrian, Hans Clusmann, Anke Hoellig, Michael Veldeman

## Abstract

**Background:** Chronic subdural hematoma (cSDH) recurrence requiring reoperation occurs in 5-33% of cases, representing a substantial clinical and economic burden. The ability to predict recurrence could enable risk-stratified surveillance protocols, potentially reducing imaging burden in low-risk patients while maintaining close monitoring for high-risk individuals. We evaluated whether machine learning algorithms could achieve clinically actionable recurrence prediction using routinely available clinical and radiographic variables.

**Methods:** This retrospective single-center study included 564 consecutive patients who underwent surgical evacuation of cSDH between 2015 and 2023. Data were randomly divided into training (75%, n=422) and test (25%, n=142) sets. We developed and compared three machine learning models—regularized logistic regression, Random Forest, and XGBoost—using 31 predictor variables including demographics, comorbidities, medications, laboratory values, hematoma characteristics, and postoperative features. Model development and hyperparameter tuning were performed exclusively on the training set using 10-fold cross-validation. The best-performing model was selected and evaluated on the held-out test set. The primary outcome was postoperative recurrence requiring reoperation.

**Results:** Postoperative recurrence occurred in 170 patients (30.1%). Within the training set, XGBoost achieved the highest cross-validated ROC AUC of 0.713 (SE=0.024), outperforming regularized logistic regression (0.686) and matching Random Forest (0.713). Variable importance analysis identified hematoma volume, coagulation parameters (INR, platelets, aPTT), and disease severity markers (ICU admission, GCS) as the most influential predictors, though absolute effect sizes remained modest. On the held-out test set, the final XGBoost model achieved ROC AUC 0.688 (95% CI: 0.590–0.772) with excellent calibration. However, at the clinically relevant 90% sensitivity threshold, test set specificity was only 30.3%, allowing potential imaging reduction in approximately one-third of non-recurrence patients. The consistency between training and test performance confirmed that limitations stem from inherent predictor information content rather than overfitting.

**Conclusions:** Machine learning models using routinely available clinical and radiographic variables cannot achieve clinically actionable risk stratification for cSDH recurrence. Despite rigorous methodology and internal validation, discriminative capacity remained insufficient to identify a low-risk patient subgroup suitable for de-escalated surveillance. These findings suggest that recurrence is driven by factors not captured in standard clinical assessment, and support either uniform surveillance protocols or symptom-driven imaging strategies rather than risk-stratified approaches.

## Introduction

Chronic subdural hematoma (cSDH) has an estimated annual incidence ranging from 7 to 30 per 100,000 individuals in population-based surveys (1–3). The incidence rises markedly with advancing age, reaching 58 per 100,000 in patients over 70 years old (4). Driven by population aging and increased use of antithrombotic medications, the global burden of cSDH continues to rise, with corresponding increases in healthcare costs and resources used (5).

The pathophysiology of cSDH involves a complex cascade of events typically believed to be initiated by minor head trauma, causing either rupture of a bridging vein or traumatic separation of the dural border cell layer (6, 7). In susceptible individuals, the initial subdural collection fails to resorb. Instead, it undergoes liquefaction and gradual expansion through a self-perpetuating cycle of inflammation, neoangiogenesis, and recurrent hemorrhage from fragile neomembranes (6, 7).

Surgical evacuation, typically performed via burr hole craniotomy with subdural drainage, remains the treatment of choice for symptomatic hematomas (8). However, postoperative recurrence necessitating reoperation occurs in 5 to 33% of cases, representing a substantial clinical and economic burden (9). Identified risk factors for recurrence include larger hematoma volume, ongoing antithrombotic treatment, history of epileptic seizures, incomplete brain re-expansion, and less organized hematoma architecture on computed tomography (CT) imaging (9, 10). Though there are good arguments in favor of only performing follow-up scans in patients with recurring symptoms (11), current practice in many centers still involves imaging for all patients following surgical evacuation. This results in cumulative healthcare expenditure which might be cause be overcarefulness.

The ability to accurately predict which patients are at highest risk for recurrence could enable risk-stratified follow-up protocols, potentially reducing unnecessary imaging in low-risk patients while maintaining close surveillance for those at high risk. However, despite numerous studies identifying individual risk factors (9), no validated clinical prediction model exists to guide personalized follow-up strategies. Traditional statistical approaches have yielded models with limited discriminative capacity, possibly reflecting the complex, non-linear relationships between clinical variables and recurrence risk that exceed the capabilities of conventional regression methods (12–15).

Machine learning algorithms, including ensemble methods such as Random Forest and gradient boosting, have demonstrated superior performance in clinical prediction tasks by capturing non-linear relationships and complex variable interactions without requiring explicit specification (16). These methods have been successfully applied to predict outcomes in various neurosurgical conditions, yet their utility for cSDH recurrence prediction remains underexplored. If sufficiently accurate, such models could identify a low-risk patient subgroup suitable for de-escalated surveillance protocols, balancing the clinical imperative to detect recurrences against the harms of repeated radiation exposure and the resulting additional costs.

The primary objective of this study was to develop and validate machine learning models to predict postoperative cSDH recurrence using variables available during the perioperative time window, incl. clinical, demographic, laboratory, and radiographic variables. Secondary objectives included identifying the most influential predictors of recurrence through variable importance analysis.

## Methods

### Study Population and Design

All consecutive patients who underwent surgical treatment for chronic subdural hematoma at a single tertiary care university hospital (RWTH Aachen University Hospital, Aachen, Germany) between January 2015 and December 2023 were considered for inclusion. This study represents an extension of previously published cohorts from the same institution (10, 17, 18), with the current analysis focusing on machine learning-based prediction of postoperative recurrence. This research was performed in accordance with all relevant guidelines and regulations, and in accordance with the Declaration of Helsinki. The study was approved by the local ethics committee (EK 25/331) and registered in the German Clinical Trials Register (DRKS00025280). Informed consent was waived due to the retrospective design. This manuscript is written in accordance with the Guidelines for Developing and Reporting Machine Learning Predictive Models in Biomedical Research (19).

Inclusion criteria comprised all patients with chronic subdural hematoma confirmed on computed tomography (CT) imaging who underwent surgical evacuation via twist drill craniostomy, burr hole craniotomy, or bone flap craniotomy. Patients were excluded if: (1) they were under 18 years of age; (2) initial CT imaging was unavailable or not uploaded to the institutional picture archiving and communication system; (3) intracranial hypotension (e.g., shunt overdrainage or spinal cerebrospinal fluid leak) contributed to hematoma formation; (4) prior neurosurgical or other cranial procedures were causally related to hematoma development; or (5) non-iatrogenic coagulation disorders (e.g., hepatogenic coagulopathy or bleeding diathesis) were present.

### Data Collection

Data were collected retrospectively through a systematic review of electronic hospital records. Extracted variables included patient demographics (age, sex), documented history of head trauma, presenting clinical symptoms and neurological deficits (paresis, gait disturbance, speech disorder, reduced level of consciousness, headache, nausea, vertigo, personality changes), Glasgow Coma Scale (GCS) score at admission, pre-existing medical comorbidities (hypertension, coronary artery disease, cardiac arrhythmias, diabetes mellitus, pulmonary disease, kidney disease, cancerous disease, prior neurological disease, alcohol abuse, prior seizure or epilepsy, peripheral arterial disease, prior deep venous thrombosis or pulmonary embolism, prior myocardial infarction or stroke, hematologic or hepatic disease), pre-admission medication use (antiplatelet therapy, dual antiplatelet therapy, oral anticoagulation including vitamin K antagonists and direct oral anticoagulants, angiotensin-converting enzyme inhibitors, angiotensin receptor blockers, statins), American Society of Anesthesiologists (ASA) physical status classification, pre-operative laboratory values (activated partial thromboplastin time, international normalized ratio, platelet count), and indication for emergency surgery.

Perioperative variables included surgical technique employed, pre-operative coagulation optimization (administration of vitamin K, prothrombin complex concentrate, thrombocyte concentrate, fresh frozen plasma, or tranexamic acid), post-operative anti-thrombotic management, use of early mobilization protocols, and post-operative need for intensive care observation. Primary outcome variables comprised of postoperative recurrence requiring reoperation after initial patients’ discharge.

### Treatment Algorithm

Surgical evacuation was indicated for patients presenting with neurological deficits (paresis, gait disturbance, speech disorder, or seizures) or for asymptomatic patients with radiographic evidence of mass effect (midline shift, ventricular compression, or sulcal effacement). An isolated headache without objective neurological findings or mass effect was not considered an indication for surgery.

The primary surgical approach was burr hole craniotomy with irrigation and placement of one or two non-suction subdural silicon drains (12-French) under general anesthesia or conscious sedation. In cases demonstrating intraoperative brain expansion with limited subdural space, a single drain was placed. Drains were typically removed after 1 to 3 days once drainage ceased. Twist drill craniostomy under local anesthesia without irrigation was reserved for homogenous hematomas in medically frail patients. Bone flap craniotomy was considered when pre-operative imaging demonstrated hyperdense clot components, suggesting complete evacuation via a burr hole approach would be inadequate. Visceral membranes were opened only if encapsulated deeper hematoma compartments were suspected; otherwise, membranous structures were left intact. The choice of surgical technique was left to the operating surgeon’s discretion. No included patients received middle meningeal Artery embolization. Pre-operative antihypertensive medications, including angiotensin-converting enzyme inhibitors and angiotensin receptor blockers, were not routinely discontinued prior to surgery. Pre-operative coagulation optimization was performed as clinically indicated based on anticoagulation status and laboratory values.

### Postoperative Management and Follow-up Protocol

Postoperative care was standardized regardless of the surgical approach, with drain removal on postoperative day 2 or 3 for patients undergoing burr hole craniotomy. Routine postoperative imaging during the initial hospitalization was performed for twist drill craniostomy cases and selectively for burr hole craniotomy patients who developed new symptoms, experienced persistent pre-operative deficits, or had clinical concern for inadequate evacuation. Identification of a significant residual hematoma with mass effect triggered revision surgery during the same admission.

Patients were discharged when complete symptom resolution was achieved or when postoperative imaging demonstrated resolution of the space-occupying effect. After discharge, surveillance CT imaging was performed 14 to 28 days post-surgery and continued at 2- to 3-week intervals until radiological resolution was documented. Asymptomatic residual hematomas without mass effect were monitored with sequential imaging until either complete resolution or clinical progression occurred. Small persistent asymptomatic subdural fluid collections without mass effect in elderly patients with brain atrophy were considered resolved, and surveillance was discontinued.

### Definition of Recurrence

Postoperative recurrence was defined as an increase in volume of residual or newly formed subdural hematoma that demonstrated mass effect (midline shift, sulcal effacement, or ventricular compression) or caused new onset or recurrence of neurological symptoms (paresis, gait disturbance, speech disorder, or reduced level of consciousness) necessitating reoperation. No absolute volume threshold was applied; clinical judgment incorporating both radiographic and clinical factors determined the need for surgical revision. Hematoma recurrence occurring more than 6 months after initial treatment, with documented complete resolution in the interim, was considered new disease rather than recurrence and was excluded from analysis.

### Radiological Assessment

Pre-operative hematoma dimensions were measured on axial CT images, including maximum length along the longest axis (mm), maximum width (mm), and total volume (mL) using software-assisted three-dimensional reconstruction (Brainlab, Munich, Germany). For bilateral hematomas, measurements of the larger hematoma were used for unilateral dimensional analysis (e.g., max. width), while in addition, the total bilateral volume was calculated. The presence and extent of midline shift (mm) were documented.

Each hematoma was classified according to internal architecture on CT imaging into one of eight subtypes by two independent assessors (HH and MV), blinded to clinical outcomes, as previously described (10). The classification system comprised: (1) homogenous hypodense, appearing hypodense relative to brain parenchyma; (2) homogenous isodense, appearing isodense to brain tissue; (3) homogenous hyperdense, appearing hyperdense without recent acute trauma; (4) sedimented, with visible layering of hemoglobin sediment separated by gravity; (5) laminar, with visible hyperdense visceral or parietal membrane; (6) bridging, with countable internal membranes connecting visceral and parietal surfaces; (7) trabecular, with complex diffuse membrane architecture precluding individual membrane counting; and (8) subacute, with acute blood components admixed within chronic hematoma in the absence of recent trauma. When features of multiple subtypes coexisted, hematomas were classified according to the most organized subtype present.

For the machine learning analysis, the eight-category architectural classification was collapsed into a simplified four-category system: homogeneous (combining hypodense, isodense, and hyperdense subtypes), organized (combining laminar, bridging, and trabecular subtypes), sedimented, and subacute.

### Data preparation and missing data handling

All statistical analyses and visualizations were conducted using R (v4.4.0; www.r-project.org) within RStudio (v2024.12.0+467). The tidyverse was used for data processing along the tidymodels framework for model development and evaluation.

Of the 630 patients in the initial cohort, 564 patients (89.5%) had complete data on the primary outcome variable (recurrence status) and were included in the analysis. Baseline characteristics were compared between included and excluded patients to assess potential selection bias. An overview of these analyses is presented in **Table 1**. The final analytic dataset comprised 31 predictor variables, including patient demographics, clinical presentation, comorbidities, medication use, laboratory values, hematoma characteristics, and postoperative features. Missing data in predictor variables ranged from 0% to 4.8%, with no single variable exceeding 5% missingness. To maximize statistical power and avoid loss of information from complete case analysis, missing predictor values were imputed using median imputation for continuous variables and mode imputation for categorical variables. Imputation was performed within each cross-validation fold to prevent data leakage.

**Table 1.**
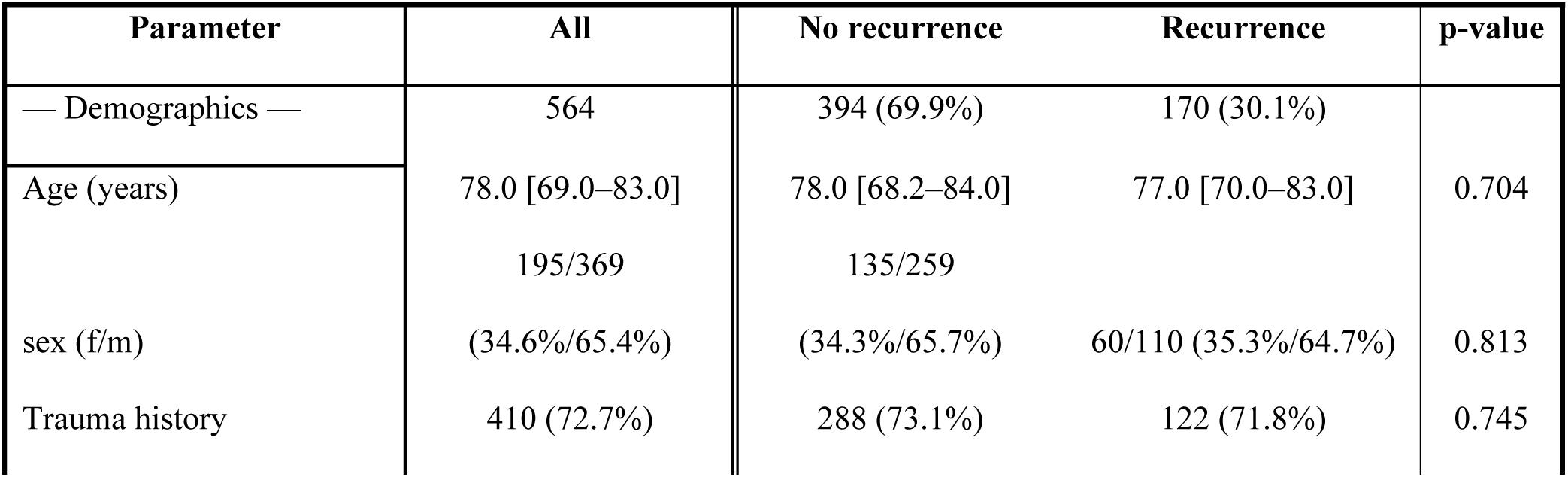
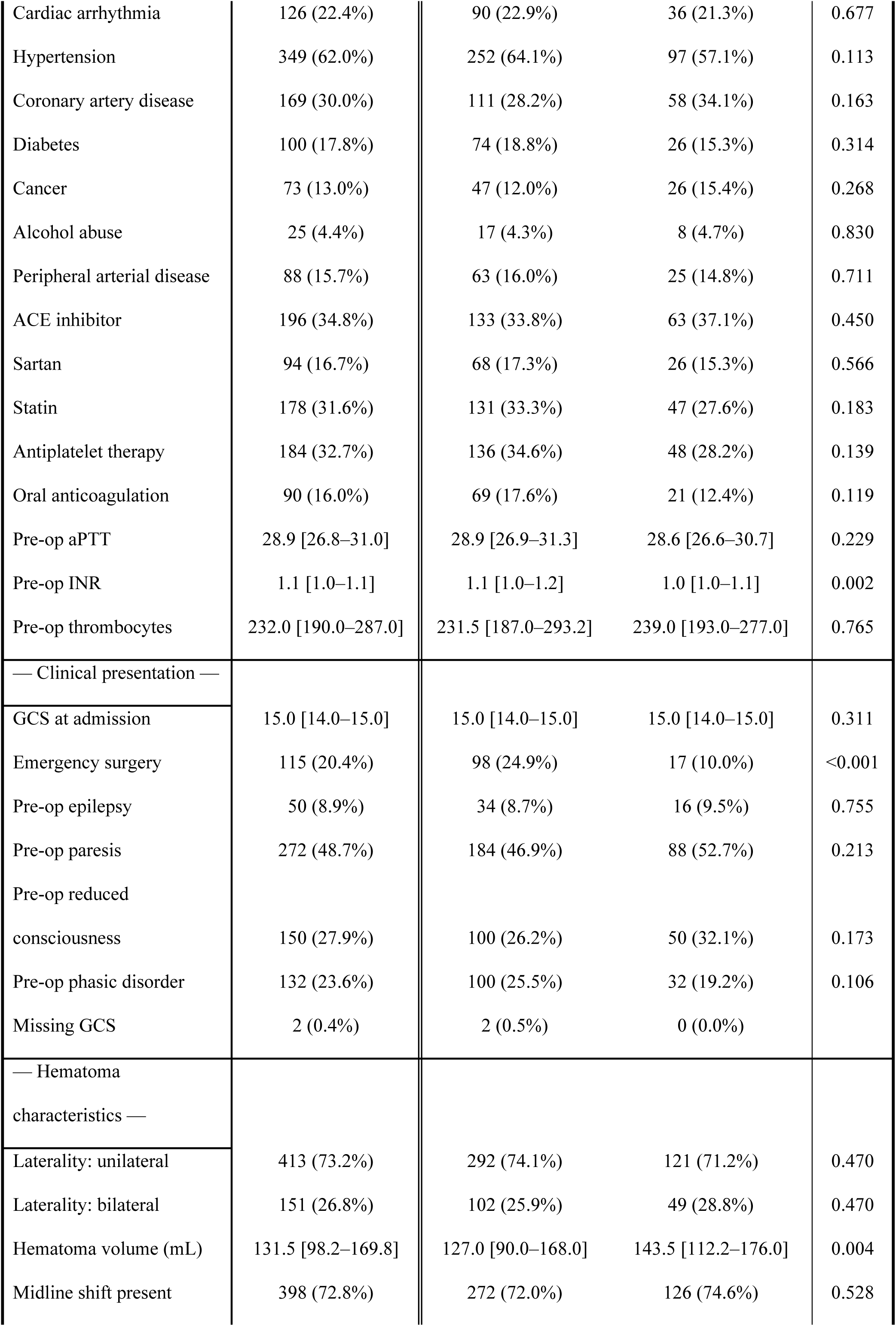
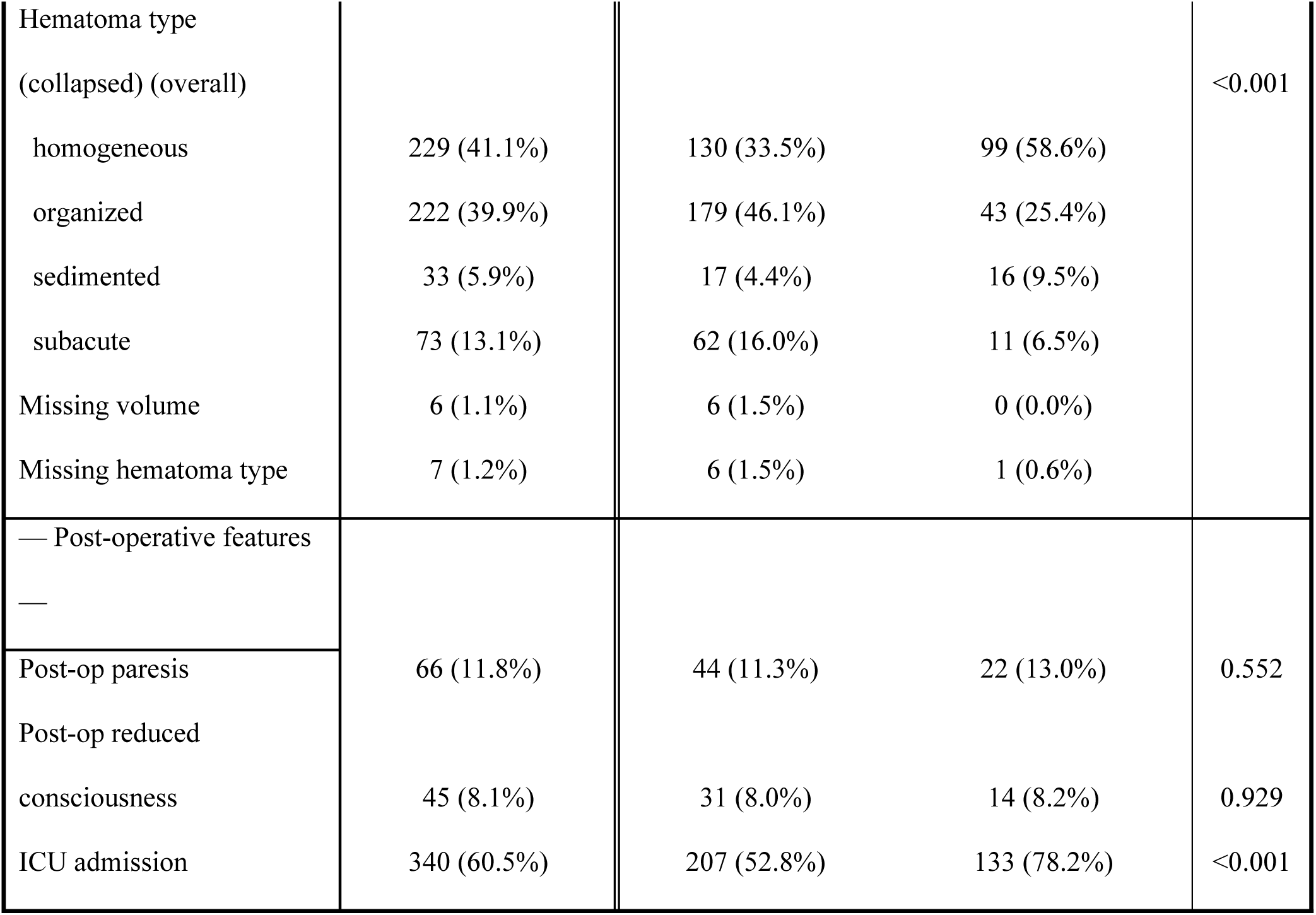
Demographic, clinical and hematoma specific parameters. Model development strategy.

The dataset was randomly partitioned into training (75%, n = 422) and test (25%, n = 142) sets using stratified sampling to maintain the outcome class distribution (approximately 30% recurrence rate) in both subsets. All model development and hyperparameter tuning were performed exclusively on the training set using 10-fold stratified cross-validation. The holdout test set was reserved for final model evaluation.

Predictor variables were preprocessed prior to model fitting. Continuous variables were standardized to have mean zero and unit variance. Categorical variables were encoded using dummy variables with reference category coding. Zero-variance predictors were identified and removed if present.

### Regularized logistic regression

We first developed a regularized logistic regression model using elastic net regularization, which combines L1 (lasso) and L2 (ridge) penalties to handle correlated predictors and perform automatic feature selection. The regularization penalty strength (lambda) and the mixing parameter (alpha, ranging from 0 for pure ridge to 1 for pure lasso) were tuned simultaneously using a grid search strategy. We evaluated 50 combinations of hyperparameters (10 penalty levels ranging from 10⁻⁴ to 10⁻⁰·⁵, and 5 mixture levels from 0 to 1) across all cross-validation folds.

### Performance metrics

Model performance was evaluated using multiple metrics appropriate for binary classification with mild class imbalance. The primary performance metric was the area under the receiver operating characteristic curve (ROC AUC). Secondary metrics included overall accuracy, sensitivity (recall), specificity, positive predictive value (precision), and negative predictive value. All metrics were calculated from cross-validation predictions and reported as means with standard errors across the 10 folds.

### Clinical decision context and threshold selection

The primary clinical objective was to identify patients at low risk of recurrence who could safely undergo less intensive follow-up. In this context, false positive predictions (classifying low-risk patients as high-risk) are clinically acceptable, as these patients would continue standard surveillance. Conversely, false negative predictions (failing to identify patients who develop recurrence) carry greater clinical risk, as these patients might be inappropriately allocated to reduced surveillance and experience delayed detection of recurrence. Therefore, model threshold selection prioritized high sensitivity (minimizing false negatives) over specificity. We considered sensitivity thresholds of 85%, 90%, and 95% as clinically relevant benchmarks, with the understanding that higher sensitivity values would necessarily result in lower specificity and fewer patients eligible for reduced imaging. The optimal threshold was selected to achieve at least 90% sensitivity while maximizing specificity, balancing the clinical imperative to detect recurrences against the potential to reduce unnecessary imaging in a subset of low-risk patients.

### Random Forest and XGBoost Models

To explore whether ensemble machine learning methods could improve upon the performance of regularized logistic regression, we developed Random Forest and XGBoost models. We constructed a Random Forest model with 500 trees and tuned two key hyperparameters: the number of predictors randomly sampled at each split (mtry, evaluated at 6 levels) and the minimum node size (min_n, evaluated at 5 levels). Hyperparameter optimization was performed using a regular grid search across all 30 possible combinations (6 × 5) within the 10-fold stratified cross-validation framework. For each hyperparameter combination, model performance was assessed across all 10 folds, and the configuration yielding the highest mean ROC AUC was selected as optimal. Permutation-based variable importance was calculated on the final model by randomly shuffling each predictor variable and measuring the resulting decrease in ROC AUC, averaged across all cross-validation folds.

XGBoost was implemented as an alternative ensemble approach. Categorical predictors were dummy-coded prior to model fitting. We trained XGBoost models with 500 boosting rounds and tuned three hyperparameters: maximum tree depth, learning rate (step size shrinkage), and minimum node size. A grid of 36 hyperparameter combinations was evaluated using the same cross-validation framework, with model performance assessed using identical metrics to enable direct comparison across all modeling approaches.

Both ensemble methods used the same imputation strategy as logistic regression, with median imputation for continuous variables and mode imputation for categorical variables applied within each cross-validation fold. Performance was compared across all three modeling approaches using ROC AUC as the primary metric, with clinical utility evaluated through threshold analysis at sensitivity targets of 85%, 90%, and 95%.

### Variable Importance and Model Interpretability Analysis

To understand which features contributed most to recurrence prediction and how they influenced model predictions, we conducted comprehensive interpretability analyses on the best-performing model. Variable importance was quantified using permutation-based importance scores, which measure the decrease in model performance when each feature’s values are randomly shuffled. This approach provides a model-agnostic assessment of feature relevance that accounts for potential interactions.

To elucidate the direction and magnitude of individual feature effects, we computed SHapley Additive exPlanations (SHAP) values. We calculated SHAP values for all features using 50 Monte Carlo samples per observation to balance computational feasibility with estimate stability. SHAP summary plots were generated to visualize both the magnitude of feature effects (mean absolute SHAP value) and the directionality of these effects as a function of feature values.

Partial dependence plots were constructed for the top continuous predictors to illustrate the marginal effect of each feature on predicted recurrence probability while averaging over the effects of all other features.

### Univariate statistics

Descriptive data are presented as mean and standard deviation (SD) for normally and as median and interquartile range (IQR: Q_1_ to Q_3_) for nonnormally distributed continuous variables. Categorical data are provided as proportions (%). After normality testing by means of the Shapiro-Wilk test, the appropriate statistical test was selected. For normally distributed continuous data, the unpaired t-Test and for nonnormally distributed data the Mann-Whitney U-Test was applied. For nominal data, the χ^2^ - Test was used. An alpha level was fixed at 0.05.

## Results

### Study Population

Between January 2015 and December 2023, a total of 630 consecutive patients underwent surgical treatment for chronic subdural hematoma at our institution. After excluding 66 patients (10.5%) with missing recurrence outcome data due to incomplete follow-up or unavailable imaging records, 564 patients (89.5%) constituted the final analytic cohort. The inclusion process is depicted as a flow-chart (**Fig. 1**).

**Figure 1.**
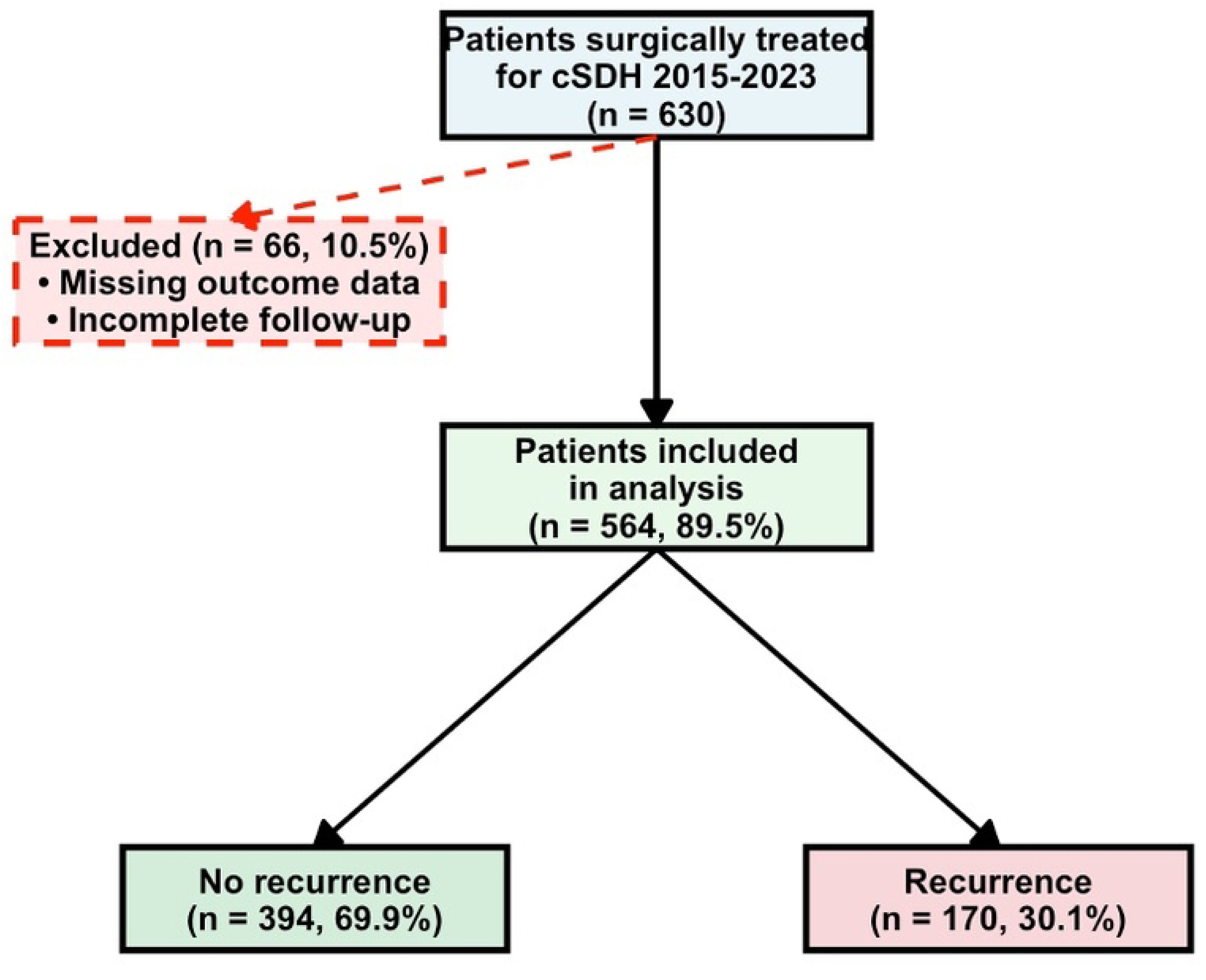
Inclusion Flow-Chart Flow diagram illustrating patient selection for the machine learning analysis of chronic subdural hematoma (cSDH) recurrence prediction. Of 630 consecutive patients who underwent surgical treatment for cSDH at a single tertiary care center between January 2015 and December 2023, 66 patients (10.5%) were excluded due to missing outcome data or incomplete follow-up, resulting in a final analytic cohort of 564 patients (89.5%). Among included patients, 170 (30.1%) experienced postoperative recurrence requiring reoperation, while 394 (69.9%) did not develop recurrence during follow-up. This class imbalance ratio of 2.32:1 (no recurrence to recurrence) was maintained in the stratified train-test split for model development and validation.

The complete cohort was randomly divided into a training set (n=422, 75%) for model development and hyperparameter tuning, and a held-out test set (n=142, 25%) for final model evaluation. The median age was 78 years (interquartile range 69-83 years), and 369 patients (65.4%) were male. A documented history of head trauma was present in 410 patients (72.7%). Pre-operative antithrombotic therapy was common, with 184 patients (32.7%) receiving antiplatelet agents and 90 patients (16.0%) receiving oral anticoagulation. The most frequently observed comorbidities included hypertension (62.0%), coronary artery disease (30.0%), and cardiac arrhythmias (22.4%).

Postoperative recurrence requiring reoperation occurred in 170 patients (30.1%), while 394 patients (69.9%) did not experience recurrence during follow-up. The recurrence rate of 30.1% represents a class imbalance ratio of 2.32:1 (no recurrence to recurrence). Baseline characteristics stratified by recurrence status are presented in Table 1. Patients who experienced recurrence had significantly larger pre-operative hematoma volumes (median 143.5 mL vs 127.0 mL, p = 0.004), lower rates of emergency surgery (10.0% vs 24.9%, p < 0.001), higher rates of ICU admission postoperatively (78.2% vs 52.8%, p < 0.001), and different distributions of hematoma architectural subtypes (p < 0.001). Homogeneous hematomas were more prevalent in the recurrence group (58.6% vs 33.5%), while organized hematomas were more common in patients without recurrence (46.1% vs 25.4%). Pre-operative INR values were slightly but significantly lower in patients who developed recurrence (1.0 vs 1.1, p = 0.002). No significant differences were observed between groups regarding age, sex, trauma history, comorbidities, or most medication use.

### Model Development and Cross-Validation Performance (Training Set)

All model development, hyperparameter tuning, and performance comparisons described in this section were conducted exclusively on the training set (n=422) using 10-fold stratified cross-validation. The held-out test set (n=142) was not used during model development and was reserved solely for final evaluation of the selected model.

### Regularized Logistic Regression Performance (Training Set)

We first developed a regularized logistic regression model using elastic net penalization to handle correlated predictors and perform automatic feature selection. Hyperparameter tuning across 50 combinations of penalty strength and mixing parameters identified an optimal configuration with a penalty of 0.053 and a mixture parameter of 0.75, favoring lasso-type regularization with substantial feature selection.

Within the training set, the model achieved a mean cross-validated ROC AUC of 0.686 (standard error 0.036 across the 10 folds), indicating modest discriminative ability (see **Fig. 2**).

**Figure 2.**
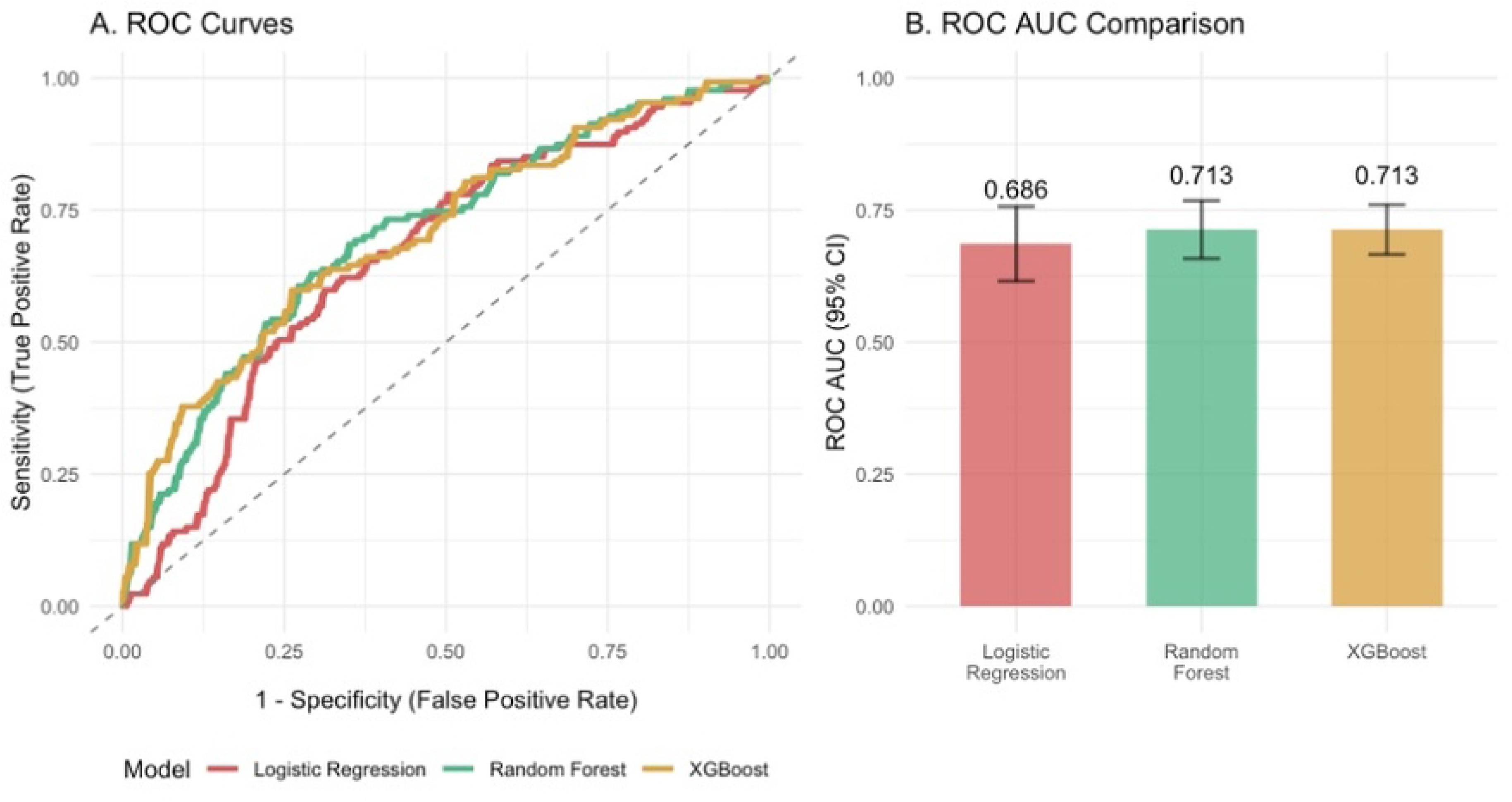
Comparison of Machine Learning Model Performance for Predicting Chronic Subdural Hematoma Recurrence **(A) Receiver Operating Characteristic (ROC) curves** comparing the discriminative performance of three machine learning algorithms: regularized logistic regression (red), Random Forest (green), and XGBoost (blue). All models were evaluated using 10-fold stratified cross-validation on the training dataset (n=422 patients). The diagonal dashed line represents the performance of a random classifier (AUC = 0.5). Curves positioned above the diagonal indicate better-than-chance discrimination between patients who developed recurrence and those who did not. **(B) Area under the ROC curve (AUC)** values with 95% confidence intervals for each model. Error bars represent 95% confidence intervals calculated from cross-validation standard errors. Both ensemble methods (Random Forest and XGBoost) achieved identical AUC values of 0.713, demonstrating modest but statistically significant improvement over regularized logistic regression (AUC = 0.686). Despite the superior discrimination metrics of ensemble approaches, all three models converged to similar performance levels, suggesting a ceiling effect determined by the inherent predictive capacity of the available clinical, demographic, laboratory, and radiographic features rather than algorithmic sophistication. The modest AUC values (0.68-0.71) indicate limited discriminative ability, corresponding to the study’s finding that these models cannot achieve clinically actionable risk stratification for surveillance imaging protocols.

However, examination of predicted probabilities revealed substantial compression of risk estimates, with all predictions ranging between 0.14 and 0.46. Patients who developed recurrence had a mean predicted probability of 0.333, compared to 0.287 for patients without recurrence. While this difference was statistically discernible and contributed to the ROC AUC of 0.686, the absolute separation between risk groups was insufficient for clinical risk stratification.

Given the clinical priority of maintaining high sensitivity to avoid missing recurrences, we evaluated model performance at sensitivity thresholds of 85%, 90%, and 95%. At all examined sensitivity thresholds, the corresponding specificity approached zero, indicating that the model could not identify any patients as low-risk without unacceptably compromising recurrence detection. Even at a sensitivity of 85%, no patients could be confidently allocated to reduced surveillance imaging. The threshold-performance curve demonstrated that achieving clinically acceptable sensitivity required classification of nearly all patients as high-risk, negating any potential reduction in imaging burden.

These findings suggest that the available baseline and early postoperative clinical variables, when combined in a linear fashion through regularized logistic regression, provide limited ability to discriminate between patients who will and will not develop recurrence.

### Random Forest Performance (Training Set)

To explore whether non-linear relationships and feature interactions could improve predictive performance, we developed a Random Forest model with 500 trees. Hyperparameter tuning across 30 combinations of predictor sampling and minimum node size identified an optimal configuration using 12 randomly sampled predictors per split and a minimum node size of 11.

Within the training set, the Random Forest model achieved a mean cross-validated ROC AUC of 0.713 (standard error 0.028 across the 10 folds), representing a modest but meaningful improvement over regularized logistic regression (delta AUC = 0.027) (see **Fig. 2**). The model demonstrated superior risk stratification compared to logistic regression, with mean predicted probabilities of 0.387 for patients who developed recurrence versus 0.276 for those who did not, yielding an absolute risk difference of 0.111 compared to 0.046 for logistic regression. Additionally, the predicted probability range expanded substantially (0.04 to 0.83) compared to the compressed distribution observed with logistic regression (0.14 to 0.46), indicating that the Random Forest algorithm successfully identified non-linear patterns and variable interactions that linear modeling could not capture.

Despite these improvements in discrimination metrics, threshold analysis revealed that the enhanced predictive performance did not translate into clinically actionable risk stratification for the intended application of reducing surveillance imaging. At a decision threshold of 0.5, the model correctly classified 274 of 295 patients without recurrence while identifying 28 of 127 patients with recurrence (sensitivity 22%, specificity 93%). However, when prioritizing high sensitivity as required for safe de-escalation of surveillance, the model’s specificity collapsed. At sensitivity thresholds of 85%, 90%, and 95%, the corresponding specificity approached zero, indicating that no patients could be confidently allocated to reduced imaging protocols without unacceptably compromising recurrence detection. This pattern mirrored the findings with logistic regression, suggesting that the limitation lies not in the modeling approach but in the inherent predictive capacity of the available clinical and radiographic features.

### XGBoost Performance (Training Set)

Within the training set, XGBoost achieved a mean cross-validated ROC AUC of 0.713 (standard error 0.024 across the 10 folds), matching the performance of Random Forest and representing a modest improvement over regularized logistic regression (see Fig. 2). Hyperparameter tuning identified optimal performance with a tree depth of 5, learning rate of 0.01, and minimum node size of 5. The model demonstrated the strongest risk discrimination among all tested approaches within the training set, with mean predicted probabilities of 0.403 for patients who developed recurrence versus 0.241 for those who did not, yielding an absolute risk difference of 0.162. The predicted probability distribution spanned the widest range observed across all models (0.04 to 0.91), and the standard error of 0.024 indicated the most stable cross-validation performance.

Despite these favorable discrimination metrics, threshold analysis within the training set revealed minimal clinical utility for risk stratification. At a sensitivity of 90%, the model achieved a specificity of only 1.6%, translating to potential imaging reduction in merely 5 of 295 patients without recurrence. This finding mirrored the results from Random Forest, confirming that the constraint on clinical utility stems from the inherent predictive capacity of the available features rather than the choice of algorithm.

Results of the clinical utility analysis and predicted probability distributions for the training set are depicted in **Fig. 3**.

**Figure 3.**
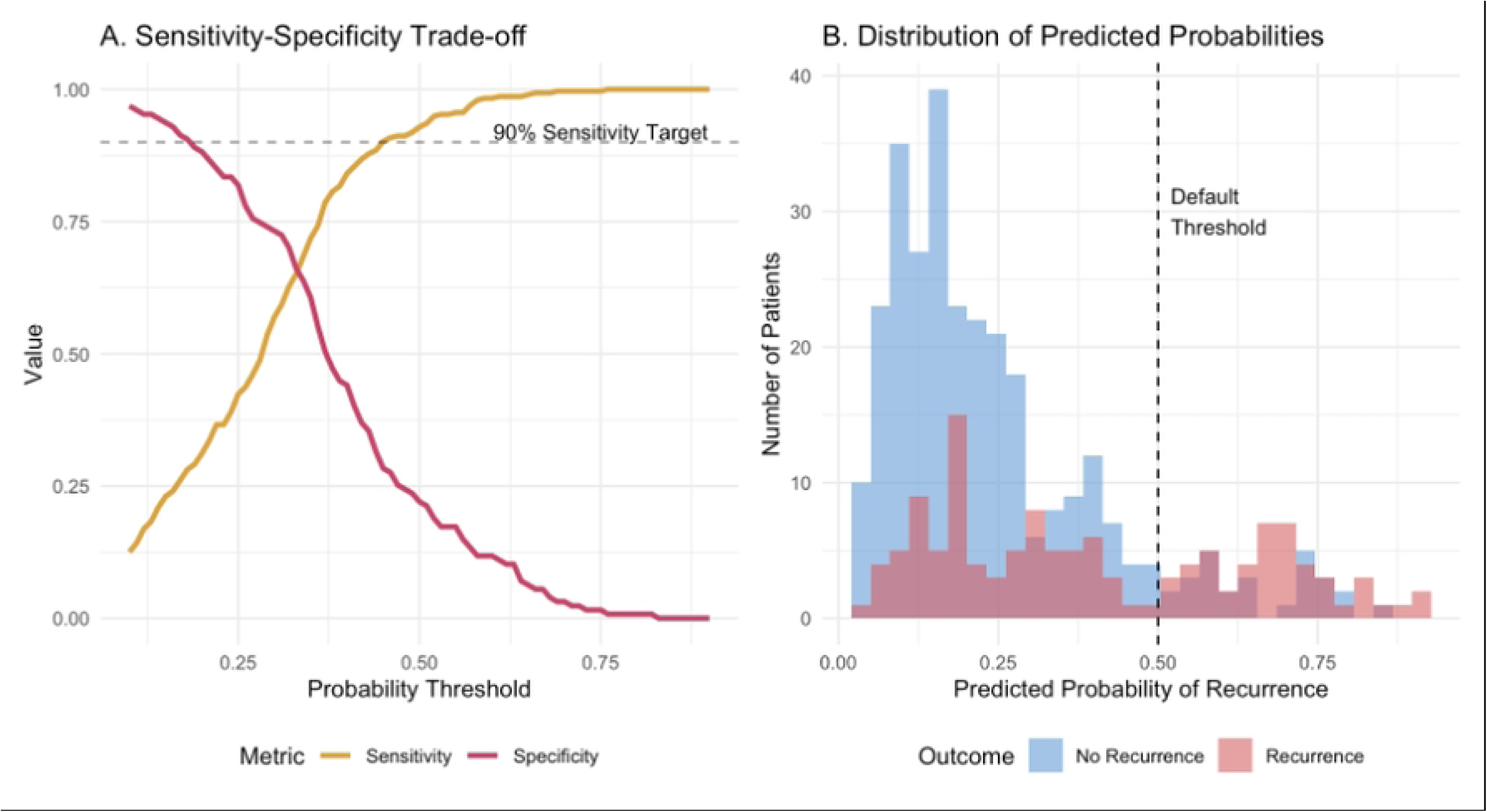
Clinical Utility Analysis and Predicted Probability Distributions. **(A) Sensitivity-Specificity Trade-off Curve** illustrating the inverse relationship between sensitivity (green) and specificity (red) across different probability thresholds for the most competitive (XGBoost) model. The horizontal dashed line indicates the clinically relevant 90% sensitivity target, representing the minimum acceptable threshold for safe de-escalation of surveillance imaging. At this sensitivity level, the model achieves near-zero specificity, demonstrating its inability to identify a meaningful low-risk patient subgroup. The steep decline in specificity as sensitivity increases reflects the compressed range of predicted probabilities and limited discriminative capacity of the model. **(B) Distribution of Predicted Probabilities** showing the overlap between patients who did not experience recurrence (yellow, n=394) and those who developed recurrence (red, n=170) based on XGBoost model predictions from 10-fold cross-validation. The vertical dashed line represents the default classification threshold of 0.5. Substantial overlap between the two distributions, particularly in the 0.2-0.4 probability range where most predictions cluster, illustrates why threshold optimization cannot achieve clinically actionable risk stratification. Only a small proportion of patients receive high-confidence predictions (probabilities >0.6 or <0.1), precluding reliable identification of patients suitable for reduced surveillance protocols. The distribution pattern provides visual evidence for the model’s limited clinical utility despite statistically significant discriminative performance (AUC 0.713).

The convergence of three fundamentally different modeling approaches (linear regression with regularization, bootstrap aggregation, and gradient boosting) to nearly identical cross-validated ROC AUC values of approximately 0.71 within the training set provides strong evidence that maximal discriminative performance has been achieved with the current predictor set.

### Rationale for Modeling Approach

We evaluated three fundamentally distinct machine learning paradigms: regularized logistic regression, Random Forest, and XGBoost. The near-identical cross-validated ROC AUC values achieved by Random Forest (0.713) and XGBoost (0.713) within the training set, combined with their similar clinical utility profiles at high sensitivity thresholds, suggested that these ensemble methods had extracted maximal predictive information from the available feature set.

Additional linear discriminant-based approaches such as LDA and QDA were not pursued, as these methods assume multivariate normality and would be expected to perform similarly to or worse than regularized logistic regression, which makes fewer distributional assumptions while accommodating the mixed continuous and categorical nature of our predictors. Naive Bayes classification was similarly excluded due to its assumption of conditional independence between predictors, which is clearly violated in clinical data where many features are inherently correlated. More complex deep learning approaches were not implemented given the modest sample size relative to the number of features and the clear performance plateau observed across the assessed approaches.

### Model Selection

Based on cross-validated performance in the training set, XGBoost was selected as the best-performing model (mean ROC AUC = 0.713, SE = 0.024), matching Random Forest performance but demonstrating superior stability (lower standard error) and the widest risk stratification (predicted probabilities ranging from 0.04 to 0.91). This model was subsequently evaluated on the held-out test set for final performance assessment.

### Variable Importance and Feature Effects (Training Set)

Permutation-based variable importance analysis conducted on the training set identified hematoma volume as the most influential predictor, accounting for 18.1% of the model’s discriminative capacity. Pre-operative coagulation parameters comprised the second tier of importance, with INR contributing 13.8%, platelet count 9.2%, and aPTT 7.4%. Disease severity markers, including post-operative need for ICU admission (8.8%) and GCS at admission (6.5%), formed the third tier, followed by patient age (6.1%) and having an organized hematoma type (7.9%). Individual comorbidities and medications showed substantially lower importance, with coronary artery disease, ACE inhibitor use, antiplatelet therapy, and statins each contributing less than 3% to model predictions. Variable importance analysis for the XGBoost model is presented in **Fig. 4A**.

**Figure 4.**
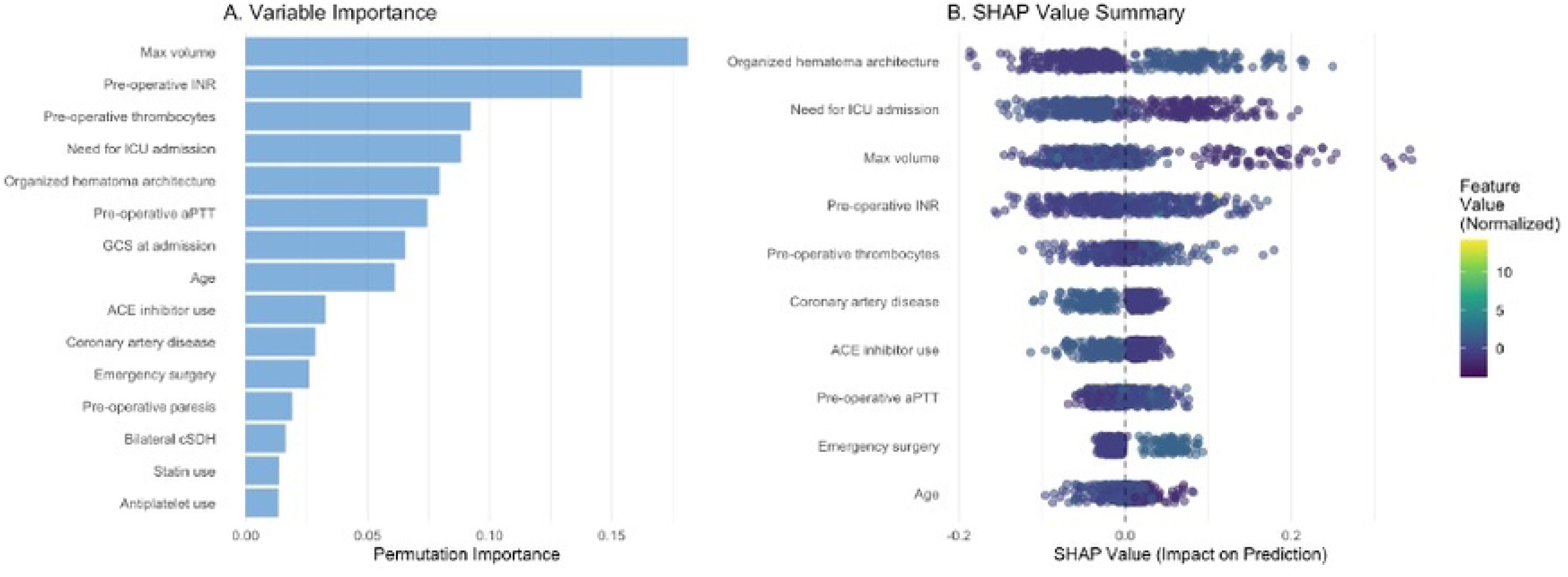
Variable Importance and SHAP Value Analysis for XGBoost Recurrence Prediction Model. **(A) Permutation-based variable importance** ranking the top 15 predictors by their contribution to model discriminative performance. Importance is quantified as the decrease in model performance when each feature’s values are randomly shuffled, with higher values indicating greater predictive relevance. Maximum hematoma volume emerged as the most influential predictor (importance 0.181), followed by pre-operative coagulation parameters (INR and thrombocytes) and disease severity markers (need for ICU admission). Individual comorbidities and medications demonstrated substantially lower importance, each contributing less than 3% to model predictions. **(B) SHAP (SHapley Additive exPlanations) value summary plot** illustrating both the magnitude and direction of feature effects on individual patient predictions. Each point represents a single patient, with position along the x-axis indicating the feature’s impact on predicted recurrence probability (positive SHAP values increase recurrence risk, negative values decrease it). Point color represents the normalized feature value (z-score) for that variable, with blue indicating below-average values and red indicating above-average values, normalized within each variable to account for different scales across features. The vertical dashed line at zero represents no effect. Despite identifying statistically important predictors, the compressed distribution of SHAP values (predominantly between-0.2 and +0.2) demonstrates that even the most influential features shift individual predictions by less than 7 percentage points from baseline. The bidirectional effects observed for several variables (points scattered on both sides of zero) suggest complex interactions between features that vary across individual patients.

SHAP value analysis revealed that despite their importance rankings, the actual magnitude of feature effects was modest. The mean absolute SHAP values for the top predictors ranged from 0.069 (organized hematoma type) to 0.019 (age), indicating that even the most influential features typically shifted individual predictions by less than 7 percentage points from the baseline. The SHAP summary plot demonstrated that higher hematoma volumes consistently increased recurrence probability (red points shifted rightward), while the effects of other top predictors showed more heterogeneous patterns.

Notably, ICU admission and organized hematoma type showed bidirectional effects across patients, suggesting complex interactions with other clinical features.

Partial dependence analysis revealed relatively flat relationships between most continuous predictors and recurrence probability across their typical ranges. Hematoma volume showed a gradual increase in predicted probability from approximately 0.28 at low volumes to 0.35 at high volumes, but without sharp threshold effects. INR, platelet count, aPTT, and age demonstrated minimal variation in predicted probability across their observed ranges. The GCS partial dependence plot showed an unexpected pattern, with predicted probabilities remaining stable around 0.65 for GCS scores of 3 through 13, then rising sharply to 0.72 for GCS scores of 14-15. This counterintuitive finding likely reflects the highly skewed distribution of GCS scores in the cohort, with 87% of patients presenting with GCS 14 or 15, and the small number of patients with lower GCS scores limiting reliable estimation of risk in that range. SHAP value analysis for the XGBoost model is presented in **Fig. 4B**.

Examination of the actual distributions of predictor variables by outcome status revealed why these features, despite their statistical importance, provided limited discriminative capacity. Mean hematoma volume differed by only 13.5 mL between patients who developed recurrence (148.2 mL) and those who did not (134.7 mL), representing a 10% relative difference. Pre-operative INR values were nearly identical (1.15 vs 1.18), as were platelet counts (237.7 vs 244.9 per μL) and patient age (75.8 vs 74.5 years). These minimal effect sizes, while statistically detectable in a dataset of this size, fall within measurement variability and lack sufficient separation to enable confident risk stratification at the individual patient level.

### Internal Validation on Held-Out Test Set

The final XGBoost model, trained on the complete training set (n=422) using optimal hyperparameters identified through cross-validation (tree depth=5, learning rate=0.01, minimum node size=5), was evaluated on the held-out test set (n=142) to assess model generalization. The model achieved a test set ROC AUC of 0.688 (95% CI: 0.590–0.772), representing a modest decrease from the cross-validated training performance (0.713, SE=0.024) that remained within confidence intervals, indicating minimal overfitting.

Test set performance metrics at the default 0.5 threshold included accuracy 73.2%, sensitivity 90.9%, and specificity 32.6%. The model demonstrated excellent calibration, with mean predicted probability (0.285) closely matching the observed recurrence rate (0.303), yielding a Brier score of 0.200. Risk discrimination in the test set remained modest, with mean predicted probabilities of 0.375 for patients who developed recurrence versus 0.245 for those who did not (absolute difference 0.130), consistent with training set findings (absolute difference 0.162). Performance of the finale modal is illustrated in **Fig. 5**.

**Figure 5.**
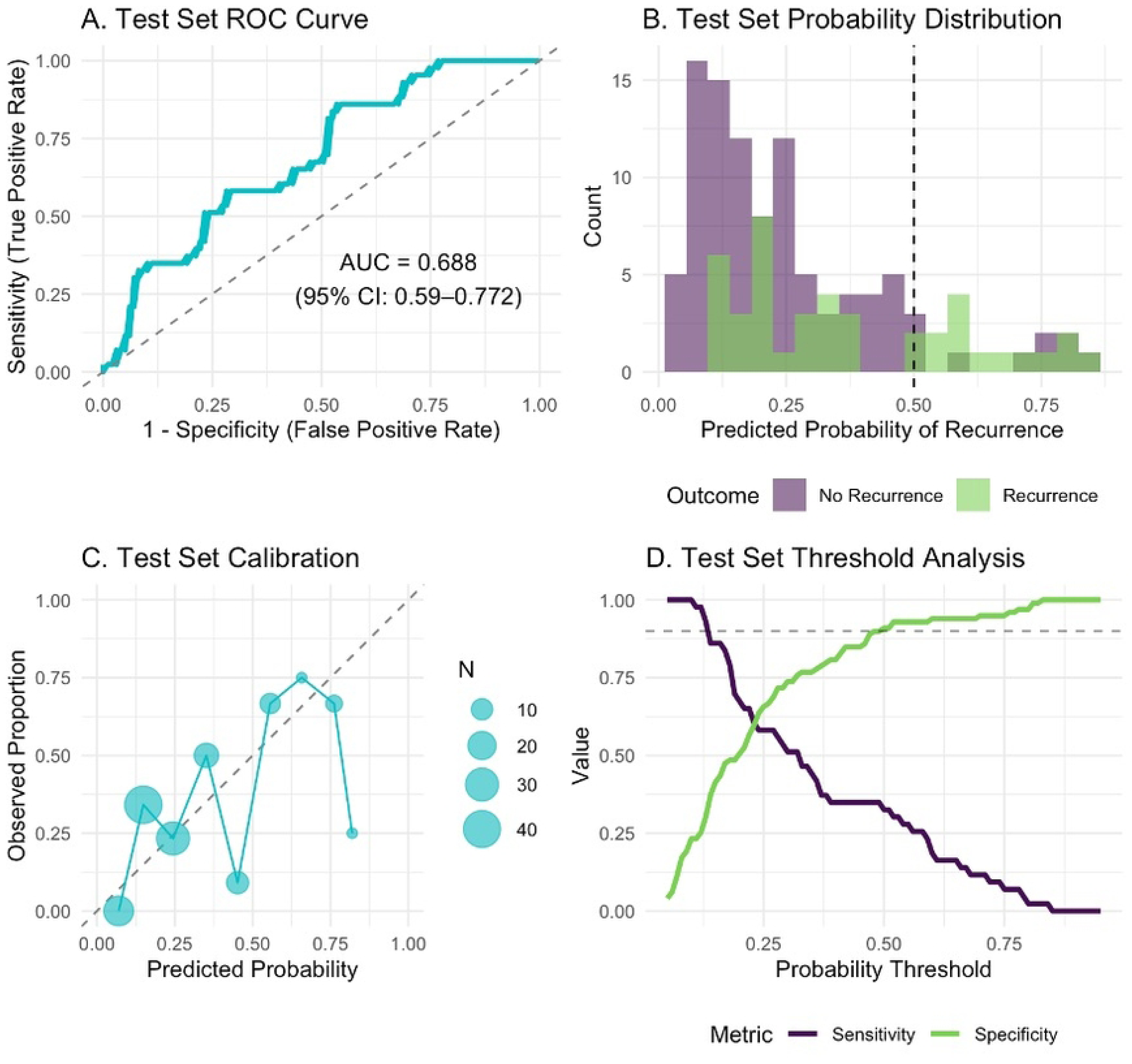
Test Set Performance and Clinical Utility Analysis of XGBoost Model **(A) Receiver Operating Characteristic (ROC) Curve.** ROC curve for the final XGBoost model evaluated on the held-out test set (n=142). The model achieved an area under the curve (AUC) of 0.688 (95% confidence interval: 0.590–0.772). The dashed diagonal line represents the performance of a random classifier (AUC = 0.5). **(B) Distribution of Predicted Probabilities.** Histogram showing the distribution of predicted recurrence probabilities stratified by actual outcome. Purple bars represent patients without recurrence (n=99); green bars represent patients with recurrence (n=43). The vertical dashed line indicates the default classification threshold of 0.5. Substantial overlap between outcome groups demonstrates limited discriminative capacity. **(C) Calibration Plot.** Agreement between predicted probabilities and observed recurrence rates across deciles of predicted risk in the test set. Points represent binned predictions, with size proportional to the number of patients in each bin (N). The dashed diagonal line represents perfect calibration. Points falling close to this line indicate good model calibration, with predicted probabilities closely matching observed outcomes. **(D) Threshold Analysis.** Sensitivity (purple line) and specificity (green line) as functions of the probability threshold for classifying patients as high-risk. The horizontal dashed line indicates the prespecified 90% sensitivity target. At this threshold (0.13), specificity was 30.3%, corresponding to potential imaging reduction in only 30 of 99 non-recurrence patients, insufficient for clinically meaningful risk stratification.

At the prespecified sensitivity threshold of 90%, the model achieved a test set specificity of 30.3%, corresponding to potential imaging reduction in 30 of 99 non-recurrence patients (30.3%). This finding validates the training set observation that current clinical and radiographic variables provide insufficient discriminative capacity for meaningful risk stratification. The consistency between training and test set performance confirms that the model’s limitations reflect the inherent information content of available predictors rather than overfitting to the training data.

## Discussion

This study systematically evaluated machine learning algorithms to predict postoperative recurrence in chronic subdural hematoma using readily available clinical, demographic, laboratory, and radiographic variables. We compared three fundamentally distinct modeling approaches, regularized logistic regression, Random Forest, and XGBoost, selecting the best-performing model for final evaluation on a held-out test set. While ensemble methods demonstrated superior risk discrimination compared to linear modeling, capturing non-linear relationships and variable interactions that logistic regression could not, none of the models achieved clinically actionable risk stratification. Threshold analysis revealed that maintaining the high sensitivity required for safe de-escalation of surveillance imaging resulted in near-zero specificity across all approaches, precluding identification of a low-risk patient subgroup suitable for reduced imaging protocols. Variable importance analysis identified hematoma volume, coagulation parameters, and disease severity markers as the most influential predictors, yet the absolute effect sizes of these features remained modest. These findings suggest that currently available baseline and early postoperative clinical features possess inherent limitations in their capacity to discriminate recurrence risk, regardless of the sophistication of the modeling approach employed.

Internal validation on the held-out test set confirmed the limited clinical utility observed during model development. The selected XGBoost model achieved a test set ROC AUC of 0.688 (95% CI: 0.590–0.772), closely matching the cross-validated training performance of 0.713, demonstrating that the model’s limitations reflect the inherent information content of available predictors rather than overfitting. The model exhibited excellent calibration in the test set, with predicted probabilities closely matching observed recurrence rates. At the prespecified 90% sensitivity threshold, test set specificity was 30.3%, allowing potential imaging reduction in approximately one-third of non-recurrence patients. While this represents some discriminative capacity, the modest specificity achieved even at this threshold remains insufficient to justify implementation of risk-stratified surveillance protocols in clinical practice. The consistency of calibration and threshold analysis between training and test cohorts strengthens the conclusion that currently measurable preoperative and early postoperative variables cannot adequately identify a low-risk patient subgroup suitable for de-escalated surveillance imaging.

### Model Performance and the Predictive Ceiling

The convergence of three fundamentally different modeling approaches to nearly identical discrimination metrics provides evidence that the observed performance ceiling reflects the inherent predictive capacity of the measured variables rather than limitations of the modeling techniques. Variable importance analysis identified hematoma volume, coagulation parameters, and disease severity markers as the most influential predictors. Examination of actual effect sizes revealed why even these algorithms could not achieve clinically useful risk stratification. The differences in these variables between patients who developed recurrence and those who did not were small, often falling within typical measurement variability. SHAP analysis quantified individual feature contributions, revealing that even the most important variables had modest effects on individual predictions.

### Comparison with Other Machine Learning Studies in cSDH

While machine learning applications in chronic subdural hematoma management are emerging, existing studies address different clinical challenges. Colasurdo et al. developed a convolutional neural network for automated detection and quantification of subdural hematomas on non-contrast CT scans, achieving high sensitivity and specificity for cSDH identification (20). Their focus on diagnostic automation differs fundamentally from recurrence prediction, serving primarily as a radiological screening tool. Fang et al. more closely aligned with our objectives by specifically predicting postoperative cSDH recurrence using a support vector machine model with combined clinical-radiomics features, reporting substantially higher accuracy and AUC than observed in our study (21). Their approach identified history of head trauma, clinical grading scores, and midline shift parameters as key predictors. However, their relatively small sample size and lack of independent test set validation may limit generalizability and could reflect optimistic performance estimates due to overfitting.

Our study’s use of rigorous cross-validation within the training set and subsequent evaluation on a held-out test set provides more conservative and likely more realistic estimates of achievable performance with standard clinical variables. The consistency between training and test performance in our cohort suggests that the modest discriminative capacity observed represents a true ceiling for prediction using routinely collected clinical and radiographic data.

### Biological and Clinical Implications

The convergence of results across modeling approaches suggests that recurrence may be largely determined by factors not captured in routine clinical assessment. Potential unmeasured determinants include molecular and genetic factors influencing inflammation and angiogenesis, microscopic characteristics of the hematoma membrane, and stochastic biological processes that are inherently unpredictable.

These findings have important implications for postoperative surveillance strategies. Our inability to identify a low-risk patient subgroup using comprehensive clinical data and machine learning algorithms, confirmed through internal validation, provides justification for maintaining uniform surveillance practices in all patients. However, this raises the fundamental question of whether routine imaging-based surveillance is the optimal approach. Schucht et al. conducted a randomized controlled trial comparing routine follow-up CT imaging versus symptom-driven imaging, demonstrating no benefit of routine surveillance on clinical outcomes (11). Their findings showed that routine imaging resulted in more reoperations and higher costs without improving patient outcomes. These results suggest that rather than attempting to refine imaging-based risk stratification protocols using machine learning, a symptom-driven approach to postoperative surveillance may be more clinically appropriate and cost-effective.

### Future Directions

Future research aimed at improving recurrence prediction will likely require identification of novel predictive features beyond standard clinical variables. Promising avenues include advanced radiomics analysis to extract subtle quantitative imaging features not appreciated by visual inspection, molecular profiling of hematoma fluid or membrane tissue to identify inflammatory or angiogenic biomarkers, and genomic or proteomic markers of individual susceptibility to recurrence. Alternatively, the consistent finding across multiple studies that recurrence prediction remains challenging may indicate that prevention strategies, such as optimization of surgical technique or pharmacological interventions targeting hematoma membrane biology, represent more promising approaches than risk stratification alone.

External validation in independent cohorts from other institutions will be essential to confirm whether the performance ceiling observed in our study represents a generalizable limitation or whether institution-specific factors influence recurrence prediction. Multi-center collaborations with larger sample sizes may also enable identification of rare but highly predictive features that cannot be detected in single-center studies.

### Limitations

This single-center, retrospective study has several limitations. First, we lacked external validation, so model performance and calibration may not generalize to other institutions, with different case mixes, and / or follow-up strategies. Second, selection and information bias are possible because 10.5% of the initial cohort lacked outcome data, and some predictors required imputation, while radiographic measurements and architecture classification are subject to interobserver and software variability. Third, the outcome definition relied on reoperation for radiographic or clinical recurrence, which may reflect local practice patterns and thresholds for revision, including our routine post-discharge imaging protocol. Finally, despite cross-validation and a held-out test set, residual overfitting and temporal drift across the 2015 to 2023 period cannot be excluded.

## Conclusion

In this single-center study, machine learning algorithms failed to achieve clinically actionable prediction of postoperative recurrence in chronic subdural hematoma using routinely available clinical, laboratory, and radiographic data. Despite employing diverse modeling paradigms, all approaches converged to a similar performance ceiling, indicating that the limitation lies in the intrinsic information content of the variables rather than in algorithmic sophistication. This highlights that recurrence is likely driven by biological, technical, or stochastic factors not captured in standard datasets. These findings support the continued practice of uniform postoperative vigilance while emphasizing that meaningful advances in risk prediction will require integration of novel biomarkers, radiomic features, or intraoperative metrics.

## Declarations

### Consent for publication

Not applicable

### Availability of supporting data

The raw data of this analysis can be made available by the authors to any qualified researcher upon reasonable request.

### Competing interests

There are no conflicts of interest to report.

### Funding

This project was made possible by the generous funding of the German Research Foundation (Deutsche Forschungsgemeinschaft, DFG), through which Michael Veldeman received a Walter Benjamin Scholarship (Grant Number: VE 1274/1-2).

### Authors’ contributions

This study was designed and conceptualized by HH, JK and MV. The manuscript was drafted by HH and MV.

Data collection was performed by HH, JK, HR, KFR, AH and MV.

HH, JK and MV were involved in the interpretation and analysis of data. This study was supervision by MV.

The final version of the manuscript was reviewed, corrected and approved by all authors.

## Data Availability

The data underlying the results presented in this study contain potentially identifiable patient information and cannot be shared publicly due to ethical and legal restrictions. Data are available from the RWTH Aachen University Hospital Institutional Data Access / Ethics Committee (contact via) for researchers who meet the criteria for access to confidential data.

## Acknowledgements

none

## Abbreviations

ACE: Angiotensin-Converting Enzyme
aPTT: Activated Partial Thromboplastin Time
ASA: American Society of Anesthesiologists
AUC: Area Under the Receiver Operating Characteristic Curve
CT: Computed Tomography
cSDH: Chronic Subdural Hematoma
DOAC: Direct Oral Anticoagulant
DRKS: Deutsches Register Klinischer Studien (German Clinical Trials Register)
EK: Ethikkommission (Ethics Committee)
GCS: Glasgow Coma Scale
ICU: Intensive Care Unit
INR: International Normalized Ratio
IQR: Interquartile Range
LASSO: Least Absolute Shrinkage and Selection Operator
NIHSS: National Institutes of Health Stroke Scale
PACS: Picture Archiving and Communication System
ROC: Receiver Operating Characteristic
SDH: Subdural Hematoma
SHAP: SHapley Additive exPlanations
STROBE: Strengthening the Reporting of Observational Studies in Epidemiology
XGBoost: Extreme Gradient Boosting

## Notes

### Competing Interest Statement

The authors have declared no competing interest.

### Author Declarations

The study was approved by the ethics committee of the RWTH Aachen University Hospital (EK20/399 & EK 25/331). Informed consent was waived due to the retrospective design.

